# Proteomic Profiling of Early-Stage Heart Failure Identifies Candidate Biomarkers and Molecular Pathways of Cardiac Inflammation in the Project Baseline Health Study

**DOI:** 10.1101/2024.12.20.24319438

**Authors:** Kalyani Kottilil, Maggie Nguyen, Megan Ramaker, Nicholas Cauwenberghs, Lydia Coulter Kwee, Sarah Short, R. Scooter Plowman, Robert J. Mentz, Pamela S. Douglas, Rury Holman, Adrian F. Hernandez, Kenneth W. Mahaffey, Francois Haddad, Svati H. Shah, the Project Baseline Health Study Group

## Abstract

**Background:** Asymptomatic structural heart disease, or stage B heart failure (HF), is clinically relevant for early HF diagnosis and prevention; circulating biomarkers could have prognostic significance for this. Thus, we performed proteomics discovery in a well-phenotyped cohort, the Project Baseline Health Study (PBHS).

**Methods:** PBHS recruited participants with and without cardiovascular risk, collecting enrollment plasma biospecimens and cardiac imaging. Proteomic profiling (N=289) was performed using mass spectrometry on 503 individuals (185 stage B HF cases, 318 stage A HF controls). Logistic regression identified stage B-associated proteins, which were then eligible for inclusion in a joint protein score derived via elastic net. Scores were assessed for incident HF prediction in the UK Biobank (UKB) and the Exenatide Study of Cardiovascular Event Lowering (EXSCEL), examined for exenatide interactions in EXSCEL, and tested for imaging trait associations in PBHS and UKB. Mechanistic analyses of multivariate significant proteins included incident HF prediction in UKB and EXSCEL, Mendelian randomization (MR), and targeted methylation loci associations with stage B HF in PBHS.

**Results:** Sixty-five proteins were associated with cases, of which 32 (49%) were retained via elastic net modeling; the resulting protein score showed good discrimination for cases versus controls (AUC 0.71, 95% CI [0.60, 0.82]). The protein score was significantly associated with incident HF in UKB and EXSCEL and left ventricular mass index (LVMI) in PBHS, as well as beneficially modified by exenatide in EXSCEL. Multivariate analysis prioritized 11 proteins associated with cases for mechanistic study, of which 4 (B2M, EFEMP1, CST3, HBB) showed significant incident HF associations in UKB and EXSCEL; CST3 and HBB were significant in MR. Methylation of cg08099136 in *PSMB8* was significantly associated with cases in PBHS.

**Conclusions:** Our findings highlight inflammatory biomarkers and mechanisms underpinning stage B HF. CpG hypomethylation at *PSMB8* may be implicated in inflammation, resulting in increased LVMI (a known HF risk factor), with circulating B2M a byproduct of this process.

**ClinicalTrials.gov Identifier:** NCT03154346

## INTRODUCTION

Heart failure (HF) is a debilitating disease with high morbidity and mortality, affecting greater than 6 million individuals in the United States (U.S.) (1). To highlight the need for early diagnosis and prevention, the American Heart Association (AHA) has recently restaged HF; stage B HF denotes the presence of structural heart disease without HF symptoms, and stage A HF is characterized by the presence of HF risk factors such as diabetes and hypertension without structural heart disease (2).

Technological advancements in molecular profiling have enabled numerous studies for identifying protein biomarkers with broad applicability to advanced HF (3–5). It is also well known that HF risk factors representative of stage A HF are drivers of structural cardiac maladaptation predictive of advanced HF; these risk factors have also been well characterized at the molecular level (6–8). However, little is known about biomarkers defining stage B HF independent of these risk factors, identification of which could uncover biomarkers of asymptomatic structural heart disease and enable earlier identification and treatment of affected individuals.

To fill this knowledge gap, we leveraged data from the Project Baseline Health Study (PBHS), a prospective population cohort study with multiple clinical and molecular endpoints, to uncover novel biomarkers and biology of stage B HF. Using echocardiography-derived HF stages and mass-spectrometry proteomics, we prioritize unique protein associations discriminative of stage B HF, independent of HF risk factors, and predictive of incident HF. We find associations of linked epigenetic loci for prioritized proteins with stage B HF to uncover inflammatory mechanisms underpinning stage B HF biology. By leveraging multi-omics data for the molecular characterization of stage B HF, we uncover putative biomarkers and biology with relevance to advanced disease prevention strategies.

## METHODS

### PBHS Study Population and Biospecimen Collection

The methodology of the PBHS has been reported previously (9). Briefly, 2,502 participants were enrolled between 2017-2019 and had extensive clinical, imaging and biospecimen assessments, and were then followed for four years with annual visits and biospecimen collection. Enrollment criteria were as follows: 40% of the cohort was selected as a general outpatient population representative of the U.S.; 60% of the cohort was selected based on high risk of cardiovascular diseases; and another 40% were selected based on high risk for oncologic disease. Clinical data necessary for HF staging were available in 2,071 of the 2,502 participants (age range, 18–92 years). Proteomic profiling was conducted in baseline enrollment plasma samples in 974 of the 2,502 participants, of which 503 were phenotyped as either stage A HF (controls, N=318) or stage B HF (cases, N=185).

### PBHS Echocardiography and HF Staging

Standard Doppler and 2D echocardiography were performed at enrollment at each site. Images were analyzed at the Duke Clinical Research Institute (DCRI) Imaging Core Laboratory, following core laboratory best practices and American Society of Echocardiography (ASE) recommendations. Echocardiographic image processing and heart failure staging in PBHS has been described (10). In brief, stage A HF controls were defined as having at least one HF risk factor but no cardiac echocardiographic abnormalities. HF risk factors included history of systemic hypertension, diabetes mellitus, obesity, cardiotoxin exposure, family history of cardiomyopathy, prevalent cardiovascular disease at enrollment, and chronic kidney disease at enrollment. Prevalent CVD was defined as coronary artery calcium (CAC) score >100 or medical history-derived coronary artery disease (CAD). HF risk factors are further described in Supplemental Table 1. Stage B HF was defined as: (1) presence of at least one HF risk factor; and (2) presence of at least one cardiac echocardiographic abnormality (left ventricular [LV] hypertrophy, LV enlargement, LA enlargement, RV enlargement, RA enlargement, LV wall motion abnormality, valvular heart disease, LV ejection fraction < 50%, RV systolic dysfunction and diastolic dysfunction). These echo abnormalities are further described in Supplemental Table 1. Participants with evidence of echocardiographic abnormalities but without HF risk factors were excluded.

### PBHS Proteomic Profiling

Untargeted, semiquantitative liquid chromatography mass-spectrometry (LC-MS) based proteomic profiling was performed in enrollment plasma samples at Verily. Briefly, mass spectra were processed using Dia-NN software and the Human PeptideAtlas (11,12). Temporal and batch effects were corrected using polynomial regression and Combat normalization (13). After quality control, 289 proteins were quantified for downstream analysis. Detailed methodology is summarized in the Supplemental Methods.

### PBHS Methylation Microarray Processing

Genomic DNA (gDNA) was extracted, quantified and bisulfite-converted from 400 ul of whole blood. Bisulfite-converted ssDNA was processed through an automated version of the Illumina Infinium MethylationEPIC microarray protocol. Resulting IDAT files were processed using minfi to generate beta, M and detection p-values for each methylation site. Detailed methodology is summarized in the Supplemental Methods.

### Statistical Analysis

The overall analytic strategy was as follows (summarized in Figure 1): levels of individual proteins were first tested for association with stage B HF vs stage A HF using univariate logistic regression models. Significantly associated proteins were then (1) used as input variables into elastic net regression models, and (2) developed into a protein score using elastic net coefficients; the protein score was (3) assessed for association with incident HF in the Exenatide Study of Cardiovascular Event Lowering (EXSCEL) and UK Biobank (UKB) cohorts; and (4) tested for association with imaging phenotypes in PBHS and UKB (details for each are below). In parallel analyses, significant proteins were tested for association with stage B vs stage A HF using multivariable models adjusted for age, self-reported race, body mass index (BMI), serum creatinine, history of hypertension (HTN), history of type II diabetes (T2D), and history of coronary artery disease (CAD). Multivariable significant proteins were further mechanistically interrogated as follows: (1) assessed for association with incident HF in EXSCEL and UKB; (2) tested for whether they were in the causal pathway for HF using Mendelian randomization; (3) used to prioritize putative CpG site methylation loci to understand regulatory mechanisms; and (4) assessed for change with exenatide, a glucagon-like peptide-1 receptor agonist (GLP-1 RA), in the EXSCEL study.

**Figure 1.**
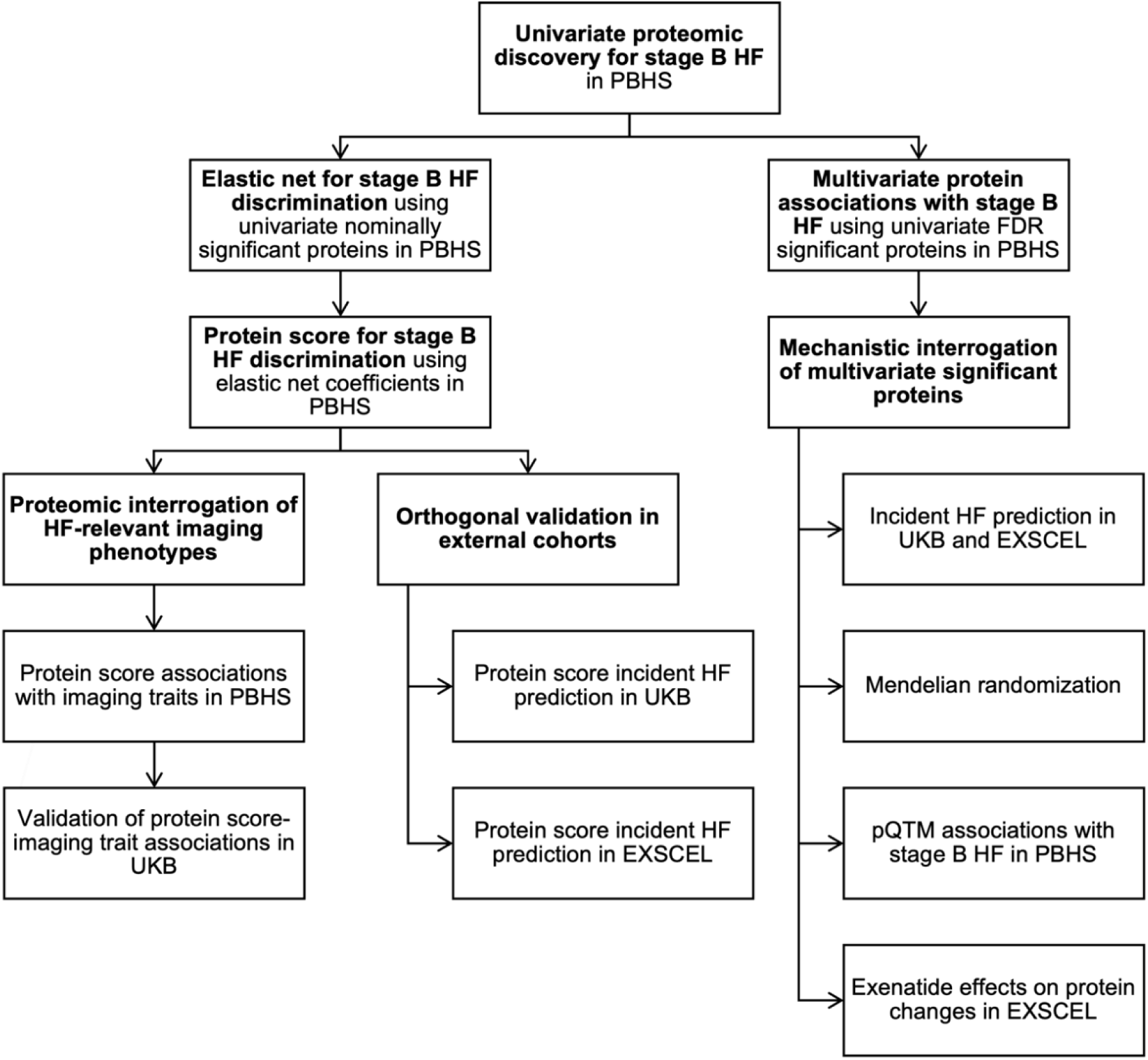
Study Overview. Overview of major analytical approaches and data sources used in the current study to identify proteins associated with stage B HF, as well as assessing their predictive ability for incident HF and mechanistically interrogating their putative functions. HF: heart failure; PBHS: Project Baseline Health Study; UKB: UK Biobank; EXCEL: Exenatide Study for Cardiovascular Event Lowering; FDR: false discovery rate; pQTM: protein quantitative trait methylation loci.

#### Penalized Regression Modeling of Significant Proteins

To quantify discrimination of stage B HF cases from stage A HF controls, an elastic net linear model was derived using nominally significant proteins (unadjusted p-value<0.05) from univariate logistic regression as input variables. Elastic net was performed in addition to multivariate logistic regression to derive weights for a clinically translatable summary protein score. Five-fold cross validation was performed on the training data (80% of the full cohort) to derive optimal model hyperparameters. Area under the receiver operating characteristic (AUC) was used to determine the model’s discriminative ability for stage B versus stage A HF in the test set (remaining 20% of the cohort). Regression coefficients of the proteins retained in the elastic net were used to create a summary protein score. This score was added to a clinical model (age, race, sex, BMI, systolic blood pressure, history of diabetes, history of hypertension, history of CAD, serum creatinine) using logistic regression and tested for discrimination of stage B versus stage A HF status on the test set. DeLong’s method was used to compare AUCs between the clinical model and the clinical model with the protein score added.

#### Orthogonal Validation of Multivariate Significant Proteins and Protein Score for Incident HF Prediction in EXSCEL and UK Biobank

The protein score and significant proteins from multivariate analyses were assessed for incident HF predictive ability in EXSCEL (14) and UKB (15). The EXSCEL study assessed the long-term cardiovascular safety and efficacy of once-weekly exenatide in patients with type 2 diabetes. In EXSCEL, HF outcomes were defined using inpatient diagnosis codes (N=129 incident HF cases, 4318 controls with no HF events). UKB is a large prospective cohort study with environmental, lifestyle, and genetic data on half a million participants aged 40-69 when recruited. In UKB, self-reported, primary care, inpatient, and death record-derived diagnosis data were used to define HF outcomes (N=1683 incident HF cases, N=36,322 controls with no HF events). Protein scores were calculated by summing the product of prioritized protein levels and their respective elastic net coefficients (from the PBHS elastic net model) for proteins that were available in each cohort. Survival analysis was performed using time-to-event Cox proportional hazards models adjusted for the same covariates in PBHS. In sensitivity analyses, we adjusted for NT-proBNP, measured in UKB and EXSCEL with Olink Explore and Somascan, respectively.

#### Imaging Phenotype Association Testing with Significant Proteins and Protein Score

Elastic net prioritized proteins and the corresponding protein score were tested for association with 12 imaging traits (Supplemental Table 2) using linear regression. Imaging variables were derived from baseline echocardiograms in the PBHS and indexed using body surface area. Proteins available in the UKB imaging sub-study were also assessed for overlapping imaging traits (Supplemental Table 2) derived from cardiac magnetic resonance imaging (cMRI).

#### Mendelian Randomization

To assess whether proteins significant in multivariable models for association with stage B HF are in the causal pathway of HF, two sample cis-MR was performed using the TwoSampleMR R package (16). Clumping distance cutoff was set to 200 kilobases (kb) to select for variants located within 200 base pairs upstream and downstream of protein gene transcription start and stop sites, respectively. Protein quantitative trait loci (pQTLs, i.e. genetic variants associated with protein levels) were selected at a significance threshold of p<1×10^-6^ and instrumental variables were selected using the HERMES GWAS meta-analysis (17). Inverse-variance–weighted estimation (IVW) was used to calculate MR estimates. Steiger filtering and outlier analysis were used to test for heterogeneity and pleiotropy, respectively. MR-Egger regression estimates were used to assess horizonal pleiotropy. Variants with heterogenous effects on HF were removed from analysis.

#### Exenatide Effects on the Protein Score and Significant Proteins from Multivariate Analysis

To assess whether the protein score and multivariate significant proteins were modified differentially by treatment with a GLP-1 RA (exenatide), linear mixed effect models inclusive of terms for treatment arm (exenatide versus placebo), protein timepoint (baseline, 12 months) and an interaction term (treatment * timepoint) with a random intercept for each subject in EXSCEL was used. These models were adjusted for BMI and HbA1c.

#### Targeted CpG Site Association Testing with Stage B HF

For further mechanistic interrogation related to potential regulatory mechanisms resulting in differential expression of proteins, we explored epigenetic regulation. Protein quantitative trait methylation loci (pQTMs: individual *cis- or trans-* CpG sites associated with plasma protein expression at genome-wide significance) were selected from a previously published epigenome wide association study (EWAS) (18) and assessed for association with stage B HF using multivariate logistic regression corrected for multiple testing and adjusted for age, sex, BMI, smoking status, and cell composition derived from clinical lab measurements.

Institutional Review Boards at Stanford University and Duke University approved the study, and participants gave written informed consent prior to participation. All association analyses were performed in Python (v3.10.12) or R (v4.4.1). Penalized regression models were developed using the scikit-learn package (19).

## RESULTS

### PBHS, UKB, and EXSCEL study populations

Baseline clinical characteristics of the PBHS study population are presented in Table 1 and baseline characteristics of the UKB and EXSCEL cohorts are presented in Supplemental Tables 3 and 4, respectively. As expected, stage B HF cases were older than stage A HF controls in PBHS (mean age, years 60 ± 14 vs 52 ± 15). Cases also had higher prevalence of self-reported Black/African American individuals (32% vs 24%) and female-identifying individuals (57% vs 51%). Age, history of hypertension, history of type 2 diabetes, and systolic blood pressure were higher in cases versus controls. Population characteristics of the UKB cMRI cohort are in Supplemental Table 5.

**Table 1.**
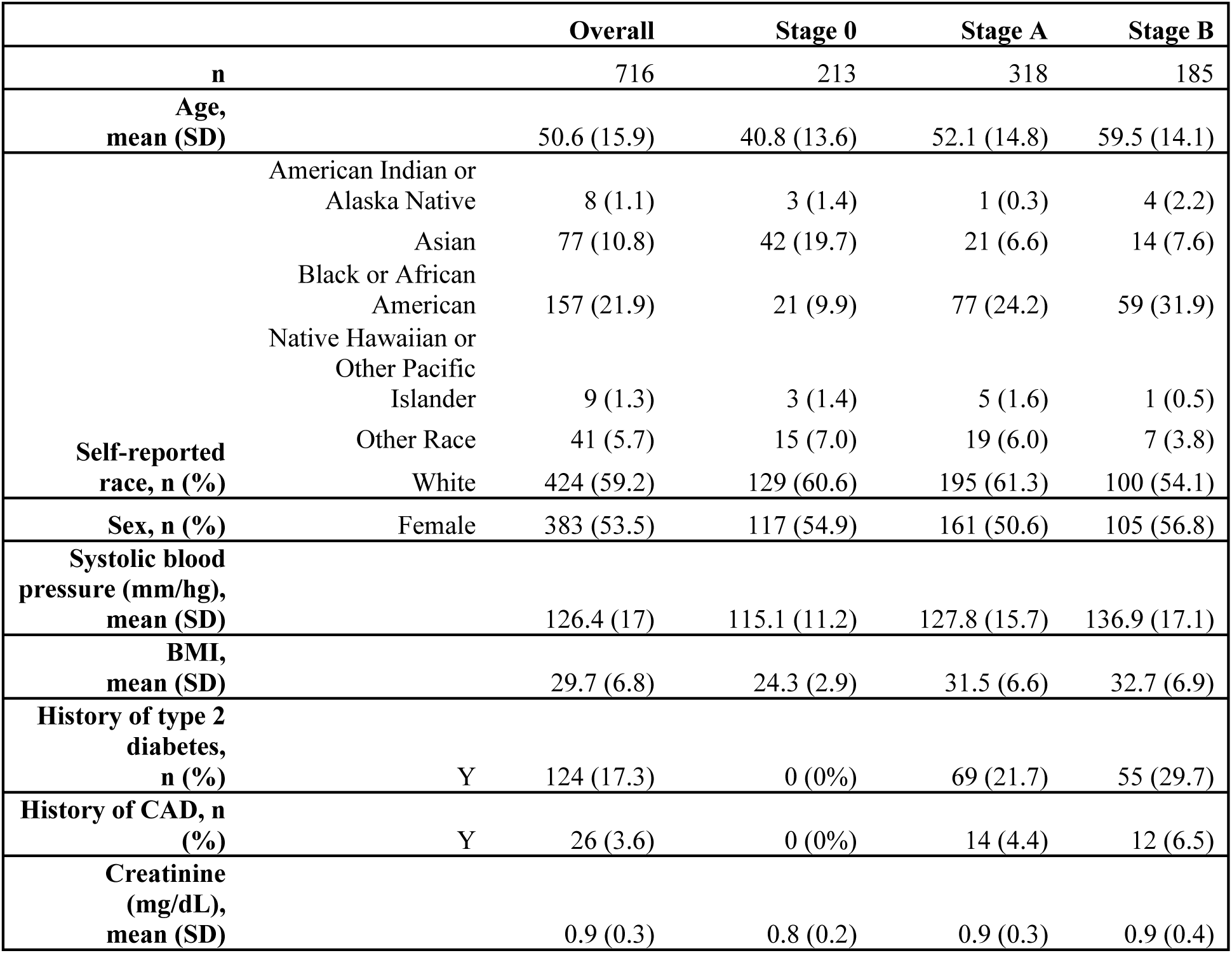
Participant characteristics by heart failure stage in the Project Baseline Health Study.

### Proteins and Protein Models for Stage B HF in PBHS

In PBHS univariate models, 65 out of 289 proteins (22%) were nominally associated with stage B compared with stage A HF (p < 0.05), and 19 were significant after FDR adjustment for multiple comparisons (Figure 2, Supplemental Table 6). Of these, 11 proteins remained associated with stage B HF in multivariable models: beta-2 microglobulin (B2M), epidermal growth factor-containing fibulin-like extracellular matrix protein 1 (EFEMP1), C-reactive protein (CRP), beta-globin locus control region (HBB-HBD-HBE1), adult beta-globin protein (HBD), cystatin C (CSTN3), hemoglobin subunit alpha (HBA1), mannosidase alpha class 1A member 1 (MAN1A1), peroxiredoxin 2 (PRDX2), serum amyloid 1 (SAA1), and serum amyloid 2 (SAA2) (p < 0.05, Supplemental Table 7).

**Figure 2.**
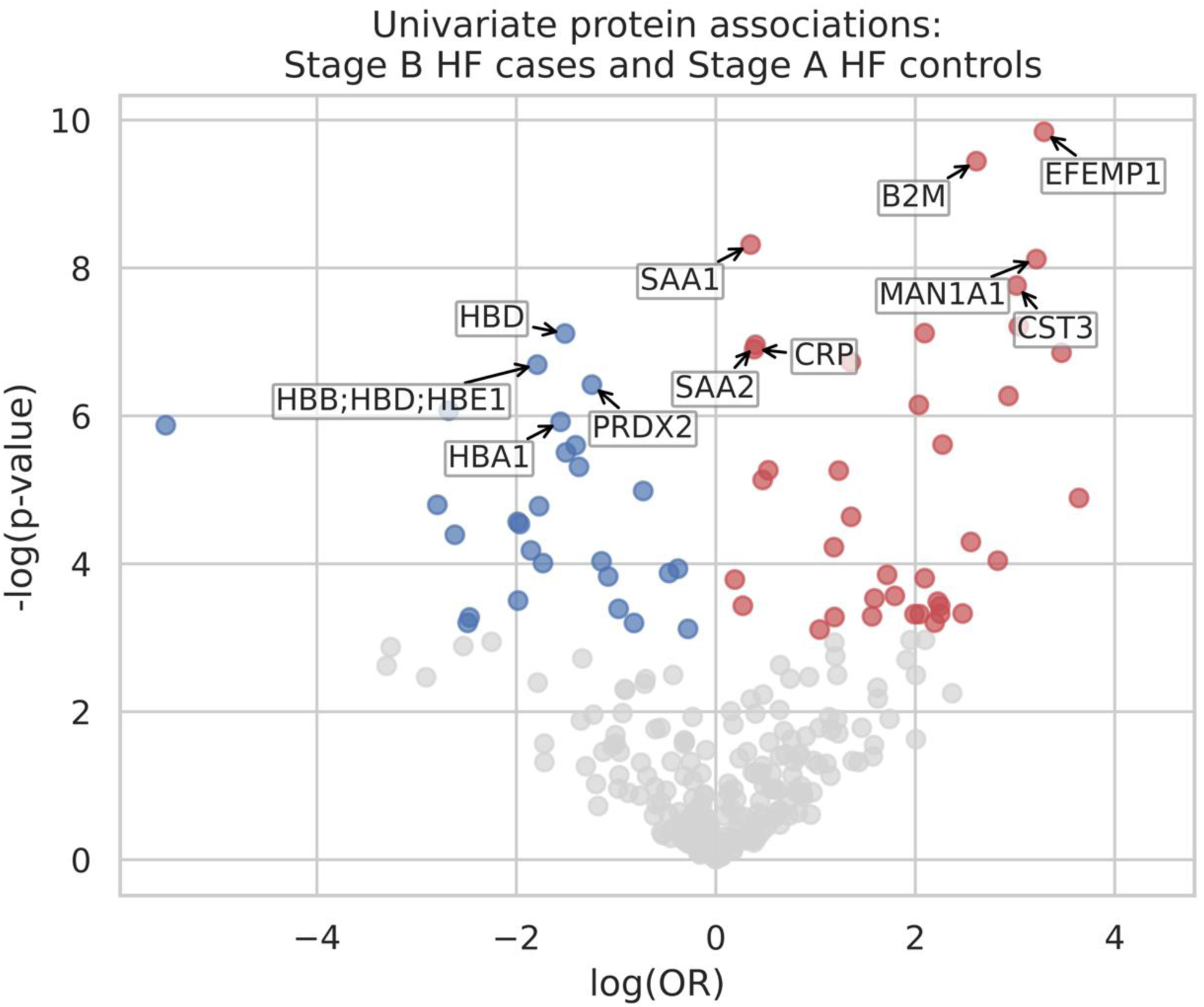
Volcano Plot Visualizing Associations of Proteins with Stage B HF. The y-axis corresponds to the −log_10_(*P*), and the x-axis corresponds to the log(OR). Univariate associations with *P* < 0.05 (not corrected for multiple comparisons) are shown in blue (if proteins were negatively associated with stage B HF) or red (if proteins were positively associated with stage B HF). Protein labels indicate proteins significant from multivariate analysis. OR, odds ratio.

We then sought to create a protein model for discriminating stage B from stage A HF. The 65 proteins nominally associated with stage B HF were used as input into elastic net logistic regression, resulting in a 32-protein model (AUC [CI]: 0.71 [0.60, 0.82], mean accuracy: 0.64). Proteins with nonzero elastic net coefficients were B2M, CRP, HBD, CSTN3, MAN1A1, SAA1, and SAA2, corroborating results from the multivariate logistic regression (Figure 3a). Other elastic net proteins are described in Supplemental Table 8.

**Figure 3.**
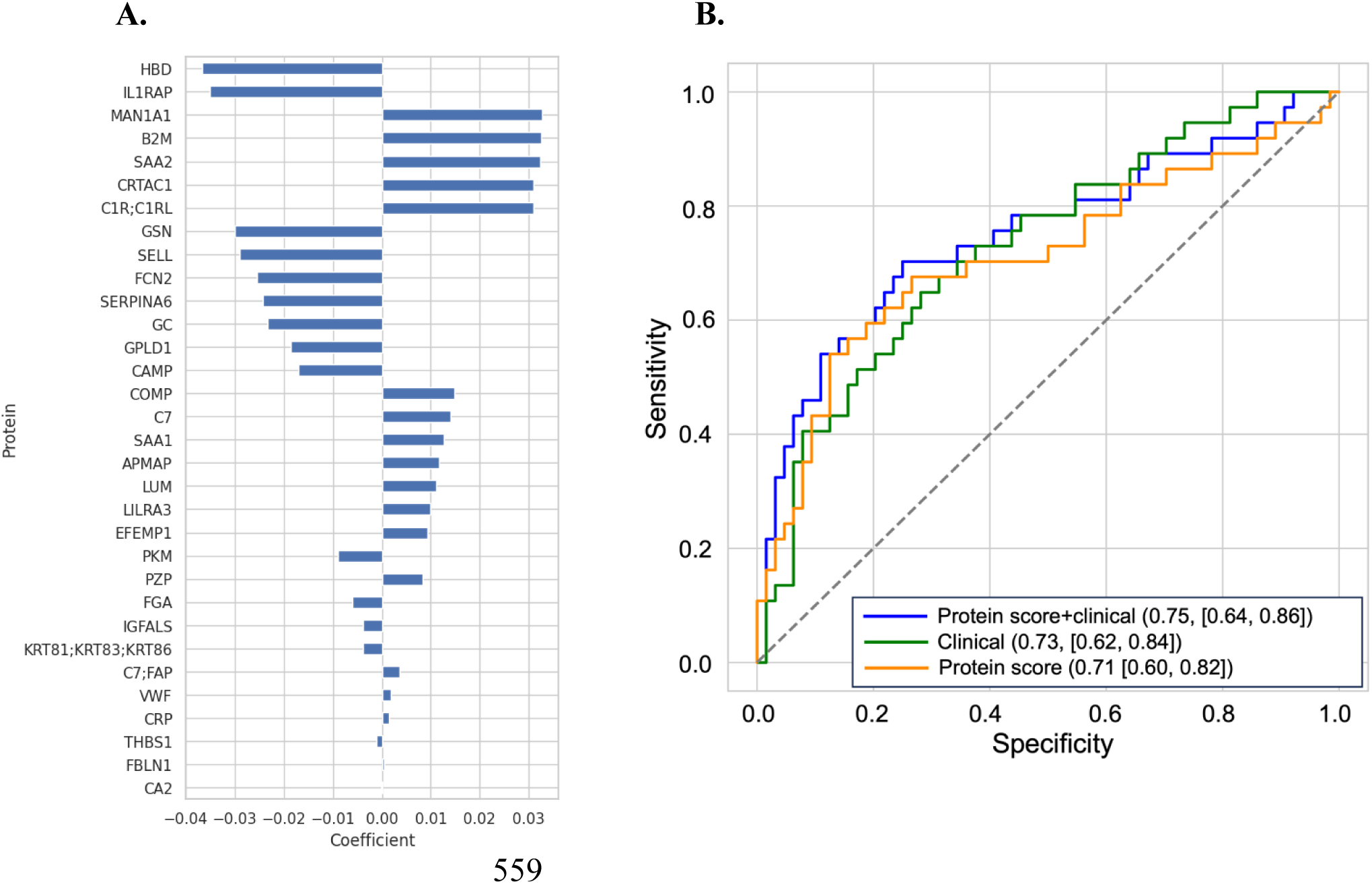
Elastic Net Derived Proteins and Protein Score for Stage B HF Discrimination in PBHS. (A) Proteins from elastic net regression for stage B HF discrimination, ordered by absolute value of coefficient. (B) Three ROC curves comparing the predictive performance of logistic regression models built with (1) protein score alone (orange curve), (2) clinical variables alone (green curve), and (3) a combined model incorporating both protein score and clinical variables (blue curve). Accuracies of the clinical, proteomic and combined risk scores in predicting stage B HF (quantified using the ROC AUC) are provided with the corresponding 95% CI. Clinical variables include age, race, sex, BMI, hypertension, systolic blood pressure, coronary artery disease, creatinine, and diabetes mellitus. ROC, receiver operator characteristic. AUC, area under the curve.

Elastic net coefficients for the 32 proteins were used to calculate a combined protein score, which performed similarly to the elastic net model for discrimination of stage B vs. stage A HF (AUC [CI]: 0.71 [0.60, 0.82], mean accuracy: 0.64). Addition of the protein score to clinical models did not significantly improve model discrimination for stage B vs. stage A HF (AUC [CI]: 0.73 [0.62, 0.84]; DeLong’s p=0.58) compared to a clinical model alone (AUC [CI]: 0.68 [0.57, 0.79], Figure 3b).

### Association of Stage B HF Proteins and Protein Score with Incident HF in the EXSCEL Clinical Trial and UK Biobank Cohort

Of the 11 proteins significantly associated with stage B HF after multivariate adjustment in PBHS, 4 (36%) were available in the UKB baseline proteomics data. Levels of B2M (p=6.7×10^-15^, HR [95% CI]: 1.14 [1.10, 1.18]), EFEMP1 (p=4.5×10^-14^, HR [95% CI]: 1.24 [1.17, 1.31]), and CST3 (p=1.9×10^-21^, HR [95% CI]: 1.34 [1.26, 1.43]) were significantly associated (after covariate adjustment) with incident HF in time-to-event multivariate Cox regression modeling in the UKB (Supplemental Table 9). Of the 32 proteins from elastic net modeling of stage B HF, 16 (50%) were available and used for protein score calculation in the UKB baseline visit. This16-protein score was also associated with incident HF after adjustment for NT-proBNP and covariates (p=2.4×10^-3^, HR [95% CI]: 1.09 [1.03, 1.15], Supplemental Table 10), but did not significantly add to a clinical model (p=0.5). Visualization using Kaplan-Meier curves by baseline protein score tertiles demonstrate higher tertiles associated with lower probabilities of freedom from incident HF (Figure 4a, Supplemental Table 11).

**Figure 4.**
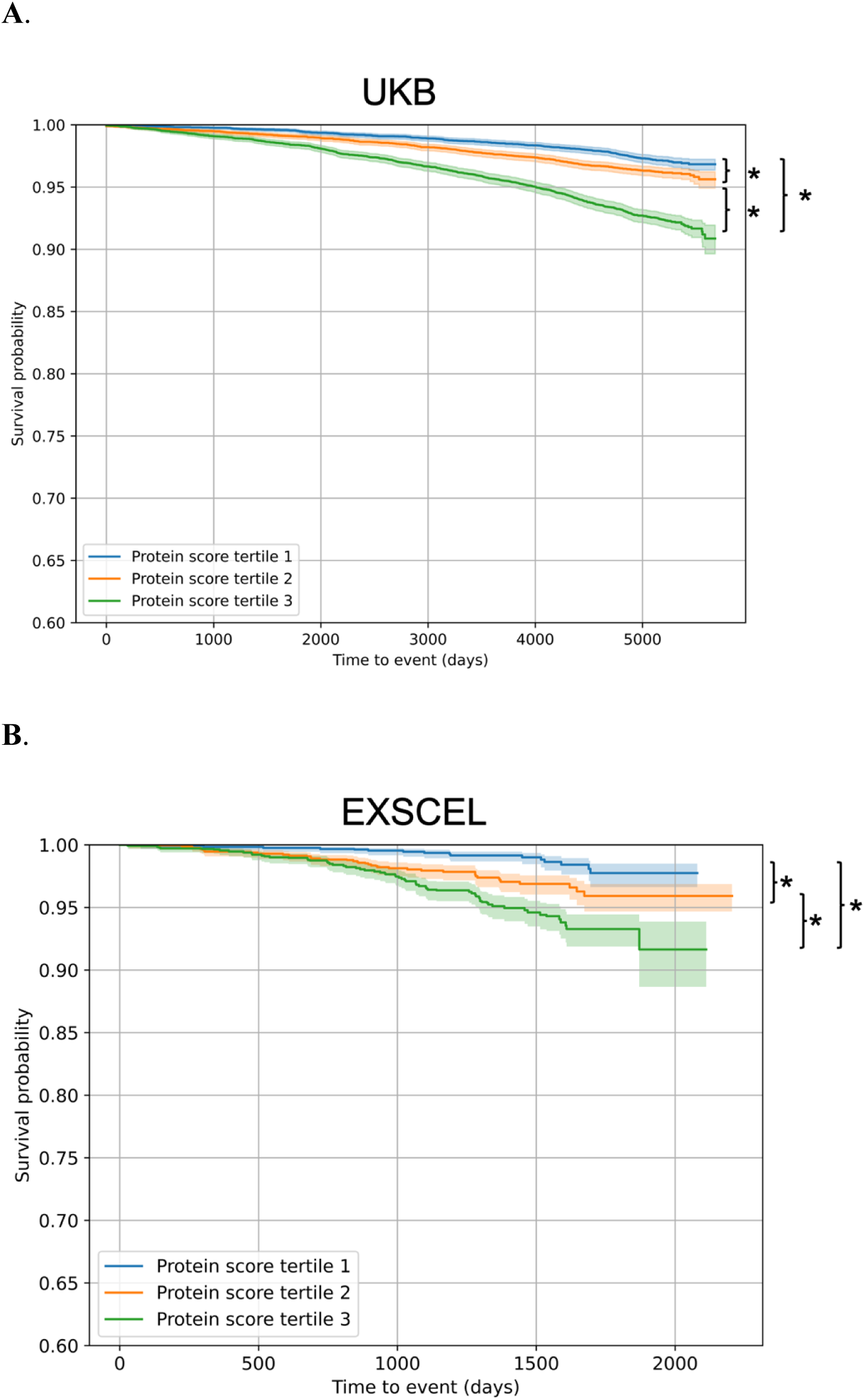
Kaplan-Meier Curves for Incident HF in UKB and EXSCEL, by Protein Score Tertiles. (A) KM curves for incident HF freedom over time in UKB, colored by protein score tertiles. (B) KM curves for incident HF freedom over time in EXSCEL, colored by protein score tertiles. The asterisk (*) represents statistically significant differences (p<0.05) in survival across the tertiles when used as a covariate in Cox proportional hazards modeling. UKB: UK Biobank; EXSCEL: Exenatide study of cardiovascular event lowering; KM: Kaplan Meier; HF: heart failure.

Nine (82%) of 11 PBHS multivariate significant proteins were measured in EXSCEL. EFEMP1 (p=9.6×10^-7^, HR [CI]: 1.62 [1.33, 1.96]), CST3 (p=5.1×10^-4^, HR [CI]: 1.1 [1.06, 1.12]), and HBD (p=7.6×10^-3^, HR [CI]: 0.79 [0.66, 0.94]) were significantly associated with incident HF in multivariate time-to-event Cox regression modeling in EXSCEL (Supplemental Table 12). Of the 32 proteins prioritized by elastic net modeling for discrimination of stage B versus stage A HF in PBHS, 25 (78%) were available in EXSCEL. The protein score was significantly predictive of incident HF in models adjusted for NT-proBNP and covariates (p-value, HR [95% CI]; EXSCEL: p=0.03, 5.78 [1.19, 28.08], Supplemental Table 13), but does not significantly improve upon a clinical model (p=0.6). Stratification of Kaplan-Meier curves by protein score tertiles demonstrate higher risk tertiles with lower probabilities of freedom from incident HF (Figure 4b, Supplemental Table 14). Of note, in linear mixed models, the protein score (p=2×10^-2^), EFEMP1 (p=1.4×10^-8^), CRP (p=1.4×10^-8^), and HBB (p=7.3×10^-2^) were found to be differentially modified by the GLP-1 RA treatment (exenatide) vs. placebo, with direction suggestive of a beneficial effect (i.e. lowering of protein levels with exenatide as compared with placebo for EFEMP1 and increasing protein levels for HBB). B2M, SAA1, SAA2, HBD, CST3, and HBA1 were not significantly modified by GLP-1 RA treatment (Figure 5, Supplemental Table 15).

**Figure 5.**
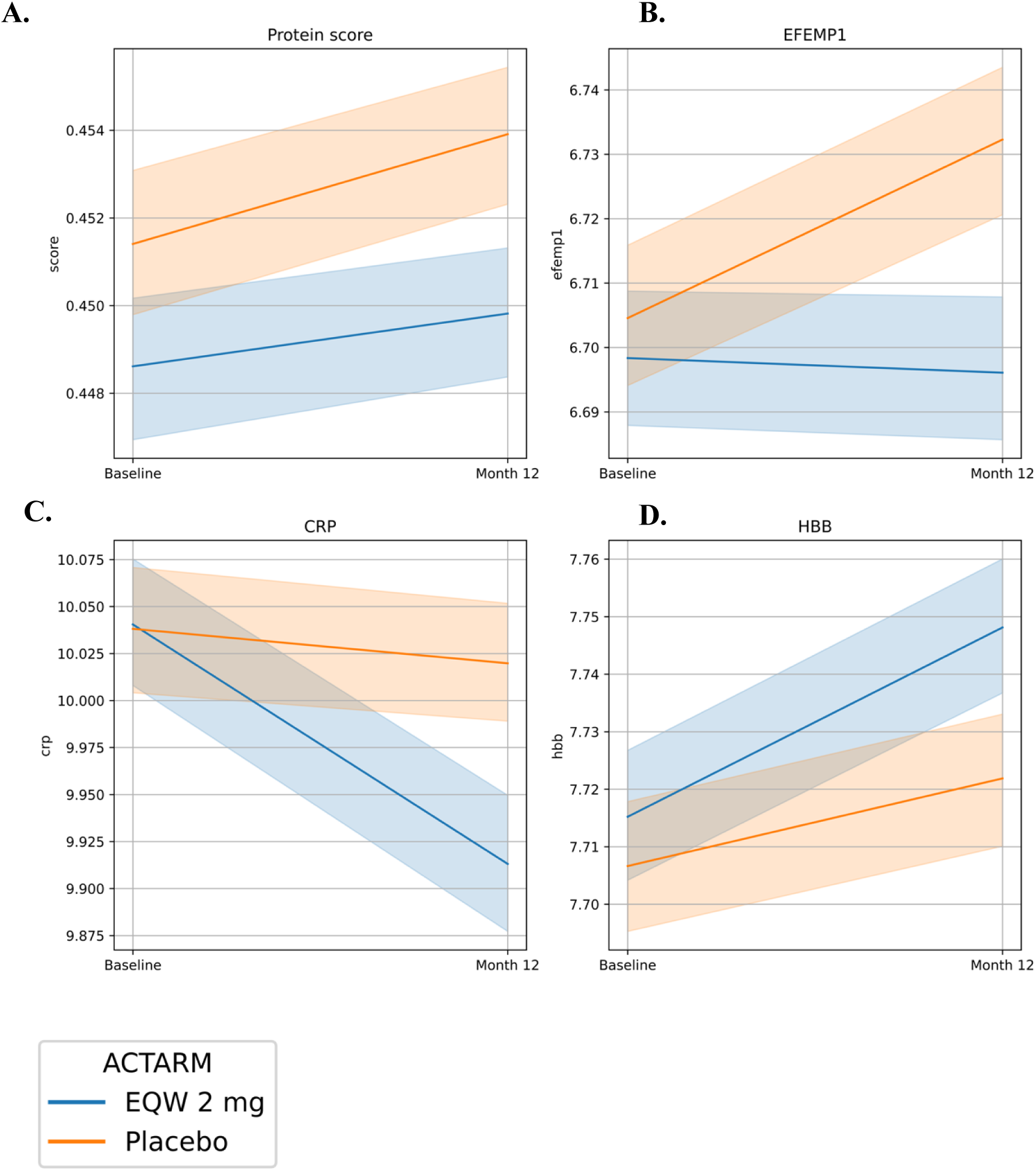
Protein Changes in Response to Exenatide Treatment in EXSCEL. (A) Changes in protein score over study timepoints, colored by treatment arm. (B) Changes in EFEMP1 over study timepoints, colored by treatment arm. (C) Changes in CRP over study timepoints, colored by treatment arm. (D) Changes in HBB over study timepoints, colored by treatment arm. EXSCEL: Exenatide study of cardiovascular event lowering; EFEMP1: EGF-containing extracellular matrix protein 1; CRP: C-reactive protein; HBB: hemoglobin subunit beta; EXSCEL: Exenatide study of cardiovascular event lowering; EFEMP1: EGF-containing extracellular matrix protein 1.

### Associations Between Proteins and Protein Score with Cardiac Imaging Traits Related to Stage B HF

In PBHS, elastic net-prioritized proteins (N=32) demonstrated overlapping associations across all imaging outcomes (N=12) in the PBHS (Figure 6a, Supplemental Table 16). The 32-protein score was associated with 4 (33%) echocardiogram-derived traits (LVRWT, LVMI, LVPWD, LAVI) after adjustment for multiple comparisons and covariates (FDR p<0.05, Figure 6b, Supplemental Table 17).

**Figure 6.**
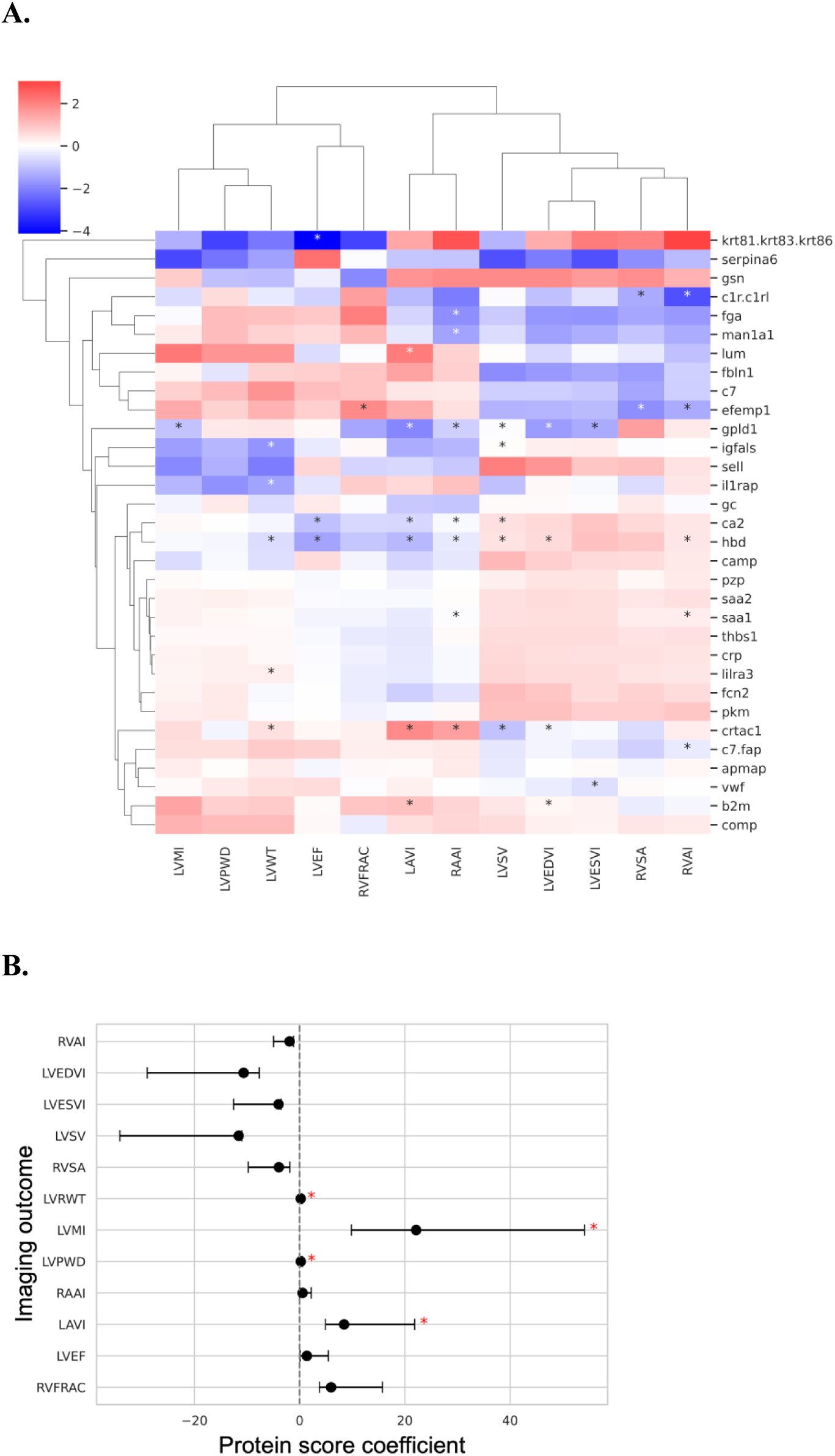
Protein and Protein Score Associations with Echocardiographic Traits in PBHS. **A.** (A) Clustered heatmap of associations between elastic net prioritized proteins and echocardiographic (echo) traits in PBHS. Heatmap is colored by linear regression coefficients. B. Forest plot showing protein score associations with echo traits in PBHS. Asterisks (*) represent significant associations after FDR and covariate adjustment. LVMI: LV mass index; LVPWD: LV posterior wall thickness in diastole; LVRWT: LV relative wall thickness; LVEF: LV ejection fraction; RVFRAC: RV fractional area change; LAVI: LA volume index; RAAI: RA area index; LVSV: LV stroke volume; LVEDVI: LV end diastolic volume index; LVESVI: LV end systolic volume index; RVSA: RV systolic area; RVAI: RV area index

Eleven (34%) overlapping proteins were measured for participants with cMRI data in the UKB. Validation of individual protein associations with similar cardiac MRI defined imaging traits in the UKB yielded generally concordant results (Supplemental Figure 1, Supplemental Table 18).

### Mendelian Randomization

Inverse-variance–weighted MR analyses supported the role of CST3, CRP, and HBA1-HBB in the causal pathway of HF and was not significant for B2M, EFEMP1, SAA1, and SAA2 (Supplemental Table 19). B2M was not along the causal pathway for LVMI.

### Epigenetic Modulation of Identified Proteins with Stage B HF and Echocardiographic Traits

To explore potential mechanisms for differential expression of our top proteins in stage B HF, we evaluated putative regulatory effects by analyzing association between protein quantitative trait methylation loci (pQTMs) with stage B HF in PBHS. Four CpG sites were identified as associated with B2M expression in a previously published EWAS (p < 5×10^-8^) (18); none were identified as associated with any other stage B HF-significant proteins. In multivariable logistic regression models adjusted for age, BMI, smoking status, and cell composition, one B2M-related pQTM (cg08099136) was associated with stage B vs. stage A HF (FDR p < 0.05, Supplemental Table 20). Linear regression derived associations between CpG site methylation and individual echocardiographic traits revealed an inverse association between cg08099136 and LVMI but was not significant after adjustment for covariates and multiple comparisons (FDR > 0.05) (Supplemental Figure 2).

## DISCUSSION

Leveraging a deeply phenotyped cohort of stage B HF, a large population study, and a large clinical trial, we demonstrate the presence of unique protein biomarkers of stage B HF independent of HF risk factors. We also show that these proteins are associated with cardiac imaging traits underlying stage B HF structural abnormalities, predict incident HF, and are beneficially modified by a GLP1-RA. These proteins reveal unexplored mechanisms of subclinical HF pathophysiology, which highlight upregulation of inflammatory pathways in asymptomatic cardiac maladaptation. Importantly, we identify proteins with potential as therapeutic targets for early disease prevention in an early and clinically important phenotype of HF not deeply studied at the molecular level.

In our study, we found higher levels of plasma B2M in PBHS participants with stage B HF as compared with stage A HF, with higher levels predicting higher risk of incident HF in EXSCEL and UKB. B2M is an important component of MHC-1 molecules and is expressed on the surface of most nucleated cells (22). Data from the Human Protein Atlas shows low expression of B2M protein in heart muscle, smooth muscle, and skeletal muscle (20). A meta-analysis of 30,988 participants showed that higher B2M levels were positively associated with CVD events even after adjustment for other inflammatory markers (albumin, CRP, and eGFR) (22). In a community-based study of 7,065 healthy individuals, B2M was associated with prevalent and incident hypertension, a known risk factor for cardiac maladaptation underpinning stage B HF (23). Notably, we demonstrated that B2M is associated with imaging traits even after adjusting for hypertension and blood pressure, suggesting that the prior associations with hypertension could be reporting on the presence or risk of structural heart disease as opposed to hypertension itself. This emphasizes the value of our study where structural heart disease was carefully phenotyped with core lab adjudicated echocardiographic phenotypes.

Other proteins found to be differentially abundant in stage B HF as compared with stage A HF with levels also predictive of incident HF were EFEMP1, CST3, and HBD. EFEMP1 and CST3 were beneficially modified by exenatide; HBB, related to HBD, was also beneficially modified. EFEMP1 was found to be upregulated in stage B HF and is important in preserving extracellular matrix (ECM) structure and cell signaling. It has been shown that EFEMP1 deletion in mouse models resulted in adverse outcomes and increased inflammation post MI, suggesting an integral role of EFEMP1 in maintaining cardiac structural integrity following ischemia (26). EFEMP1 protein expression has also been found to be two-fold higher in left ventricular tissue of heart failure patients relative to non-failing patients, and is thought to maintain cardiac tissue integrity after ischemic damage caused by events such as myocardial infarction (26). CST3 is an established biomarker of kidney function and has been previously identified as a marker of inflammation and a prognostic marker for heart failure independent of creatinine or eGFR (27). Mechanistically, CST3 is implicated in elastin and collagen degradation via cysteinase inhibition, which is known to cause extracellular matrix remodeling in the development of HF (28). HBD is a subunit of hemoglobin; HBD, along with other hemoglobin subunit proteins (HBB, HBA1) were also found to be lower in stage B HF as compared with stage A HF. Low hemoglobin can be indicative of anemia, a known HF comorbidity and marker of HF severity; interestingly, HBB, another hemoglobin subunit, was found to be higher in participants who were administered exenatide, indicating the potential utility of such therapeutics in mitigating anemia-related complications of early-stage HF.

In addition to these prioritized proteins, which showed consistent effects across different analyses, we show that a protein score for stage B HF is beneficially modified by exenatide and predictive of incident HF in two external cohorts, even after adjustment for NT-proBNP. Further investigation of the protein score’s relationships with structural cardiac features reveals LVMI to be the most strongly associated with the protein score and many of the individual proteins. LVMI is a strong subclinical risk marker for left ventricular hypertrophy (LVH), which increases risk for heart failure – association of the protein score, and significant proteins such as B2M, with LVMI further underscores the importance of inflammatory and fibrotic processes underlying early HF pathophysiology.

Epigenetic mechanisms may play an important role in regulating gene expression in cardiovascular disease pathophysiology. Herein we find that a protein quantitative trait methylation locus (pQTM) for one of our top proteins (B2M) was associated with stage B HF in PBHS. The CpG site (cg08099136) is located at the PSMB8 locus on chromosome 6 in an active regulatory region (29). PSMB8 encodes for a subunit of an immunoproteosome upregulated by NLRC5, which increases expression of B2M protein via MHC-1 (30). Hypomethylation of this region may lead to increased circulating expression of the immunoproteosome and immune activation (31). However, Mendelian randomization did not support the role of B2M in being in the causal pathway of HF, suggesting the observed higher levels in stage B vs. stage A HF may be related to existing structural cardiac changes. This is reflected by our observation that B2M is most associated with the cardiac imaging trait of LVMI and by known expression of B2M in cardiac tissue. Despite not being significant, the inverse association shown between cg08099136 methylation and LVMI preliminarily supports the role of this epigenetic modification in adverse cardiac structural remodeling. In summary, our work demonstrates the potential role of epigenetic mechanisms (i.e. hypomethylation at the PSMB8 locus) leading to immune activation (reflected in higher circulating and cardiac tissue-specific B2M levels) and left ventricular hypertrophy (a stage B HF defining feature) as important, early features of HF pathophysiology (Figure 7).

**Figure 7.**
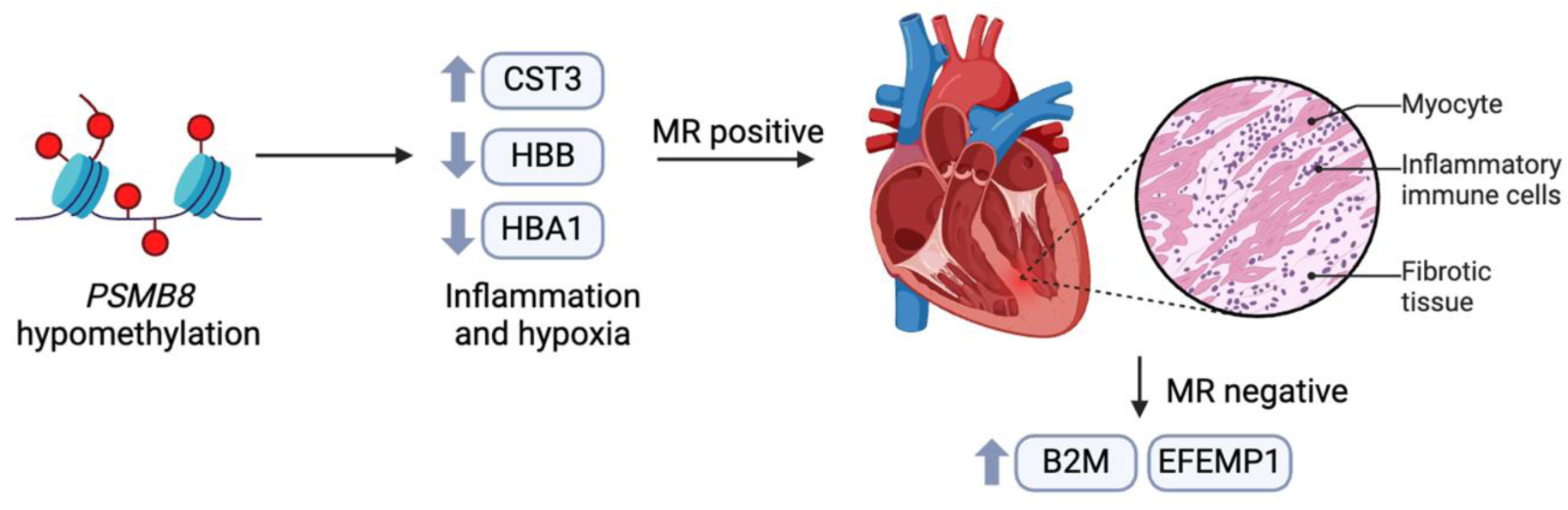
Summary of Conceptual Model. Generated in BioRender.

The strengths of this study include the focus on stage B HF, a clinically relevant phenotype of HF that has not been characterized at the molecular level, in a study with careful stage B HF phenotyping through systematic, core lab–based echocardiographic evaluations in a diverse cohort of participants. We also highlight clinical relevance with regards to the ability of these proteins to predict incident HF independent of HF risk factors in a large clinical trial and a large population-based cohort. Limitations to this study should be noted: we did not have NT-proBNP levels in PBHS, a gold standard blood protein biomarker for identifying HF with high predictive performance. However, we show that the predictive capability of our top proteins, B2M, EFEMP1, CST3, and HBB for incident HF remained significant in EXSCEL and UKB even after adjustment for NT-proBNP.

In this study, we demonstrate the utility of high throughput proteomic profiling in identifying potential biomarkers of early-stage HF in a large population study. B2M, which is shown to also be associated with stage B HF, may contribute to cardiac fibroblast activation, which in turn promotes healing and myocardial repair after ischemia. By interrogating proteins prioritized for association with early-stage HF and epigenetic regulation of these proteins, we are able to more deeply characterize inflammatory environments known to facilitate HF development.

## Data Availability

The deidentified PBHS data corresponding to this study are available upon request for the purpose of examining its reproducibility. Requests are subject to approval by PBHS governance.

## ACKNOWLEDGMENTS

The authors wish to thank Project Baseline Health Study participants and study sites.

## Ethics Statement

The study was approved by the Duke University and Stanford University Institutional Review Boards. Informed consent was obtained from all participants enrolled in the Project Baseline Health Study in accordance with the Declaration of Helsinki.

