## Supplemental Material for "Proteomic Profiling of Early-Stage Heart Failure Identifies Candidate Biomarkers and Molecular Pathways of Cardiac Inflammation in the Project Baseline Health Study"

**Supplementary Methods**

*Proteomic profiling*

Untargeted, semiquantitative liquid chromatography mass spectrometry (LC-MS)-based proteomic profiling was performed in enrollment plasma samples utilizing the proteomics pipeline at Verily, Inc., utilizing robotic liquid handling and validated plasma preparation kits. Specifically, for each plasma sample, 2 microliters were denatured with trypsin/Lys-C protease and the subsequent peptides were desalted and dried down in a vacuum concentrator. Dried pellets were dissolved in 40 microliters of 0.1% (v/v) formic acid, then peptide concentrations were normalized to 1 microgram per microliter and combined with iRT standard peptides (1:20 v/v). Five (5) micrograms of each sample were randomly injected in duplicate onto a customized microflow high-resolution liquid chromatography-mass spectrometry (LC-MS) setup. Mass spectra were acquired in data-independent acquisition (DIA) mode for accurate and reproducible quantification.

Each sample was run as two technical replicates for each batch. If instrument performance degraded during a batch, more than two replicates were processed. Mass spectra were stored as ThermoFisher raw files, which were converted to mass spectrometer output files using the msconvert function from ProteoWizard [11]. Mass spectrometer output files were centroid normalized. Dia-NN (v1.8.1), a universal software for DIA proteomics data, was executed in library-free mode to generate preliminary peptide abundances [12]. The plasma proteome from the Human PeptideAtlas [13] was used to restrict the search space to plasma protein sequences. A subset of samples was jointly analyzed to generate the spectral library with Dia-NN. The first available replicate of each sample was selected from each participant’s entry visit. Each replicate sample was reprocessed independently using the newly generated spectral library.

The latest batch was used for analysis if repeat batches were performed. Samples and precursors were included if: replicates had more than 2500 precursors; samples had at least two valid technical replicates; proteins had a Dia-NN library q-value less than 0.01; proteins had Dia-NN q-value less than 0.05 in both replicates across at least 100 samples; precursors had coefficients of variation less than 0.2 for all samples and were reproducible between replicates. Microbial proteins, contaminants and lg variable chain proteins were excluded from downstream analysis.

Polynomial regression for log-transformed precursor expression was fit on the run order for each precursor in each batch, and predictions were regressed out to the median to adjust for temporal bias. Non-log transformed normalized precursor quantities were summed up to compute protein abundances within each replicate. Protein quantities were log-transformed again and averaged between technical replicates to obtain protein quantities at the sample level. Missing values in one replicate were imputed with values from the other replicate. Averaged protein quantities were corrected for batch effects using a Python implementation of the combat method [14]. Filtering was performed to select for participants with proteomics data at the baseline PBHS visit, across each of which quantification measurements for 289 proteins remained after quality control.

*Methylation array*

Samples were processed over the span of three years in-house at Verily Life Sciences. The first 100 y0/study start samples were processed in 2017, 1,451 additional Y0/study start samples were selected based on specimen availability at the time and processed in 2018, and 488 (inclusive of longitudinal timepoints and intentionally selected for the “Diabetes of the Immune System” and Biomarkers of Organ Injury” sub-studies) were processed in 2019. The total number of samples processed to date is 2,039, from a total of 1,751 unique participants.

Genome DNA (gGNA) was extracted from 400 uL of previously frozen K2/EDTA whole blood stored in 1 mL aliquots using the QI symphony DSP DNA Midi Kit (96) (QIAGEN, Hilden, Germany) run on the QIAsymphony SP (QIAGEN, Hilden, Germany). Processing was performed in batches of 24 samples per run, including a negative control of molecular grade water. Extracted gDNA was quantified using the Quant-iT PicoGreen dsDNA Assay Kit (Invitrogen, Waltham, MA) and run on a CLARIOstar microplate reader (BMG LABTECH, Ortenberg, Germany).

The resulting bisulfite-converted ssDNA was normalized to an input amount of 400 ng in a volume of 20 uL and arrayed into a 96-well plate format with up to 93 patient samples per plate. Human universally methylated and non-methylated controls (Zymo Research, Irvine, CA, USA) as well as gDNA from a control cell line (Coriell Intitute for Medical Research, Camden, NJ, USA) were added to the remaining 3 well positions on each plate. Samples underwent automated bisulfite conversion, desulphonation, bead-based cleanup, and elution using an EZ-96 DNA Methylation-Lightning MagPrep kit (Zymo Research, Irvine, CA, USA) per the manufacturer’s protocol. Four uL of bisulfite-converted ssDNA for each sample and control was processed through an automated version of the standard Illumina Infinium MethylationEPIC microarray assay protocol (Illumina Inc., San Diego, CA, USA) inclusive of amplification, fragmentation, precipitation, resuspension, hybridization to BeadChips, washing, and extension and staining (XStain). BeadChips were scanned on an Illumina iScan (Illumina Inc., San Diego, CA, USA) which generated IDAT files for downstream computational processing.

The resulting IDAT files were computationally processed using minfi with background normalization (Noob) and dye correction to generate beta-, M-, and detection p-values for each site.

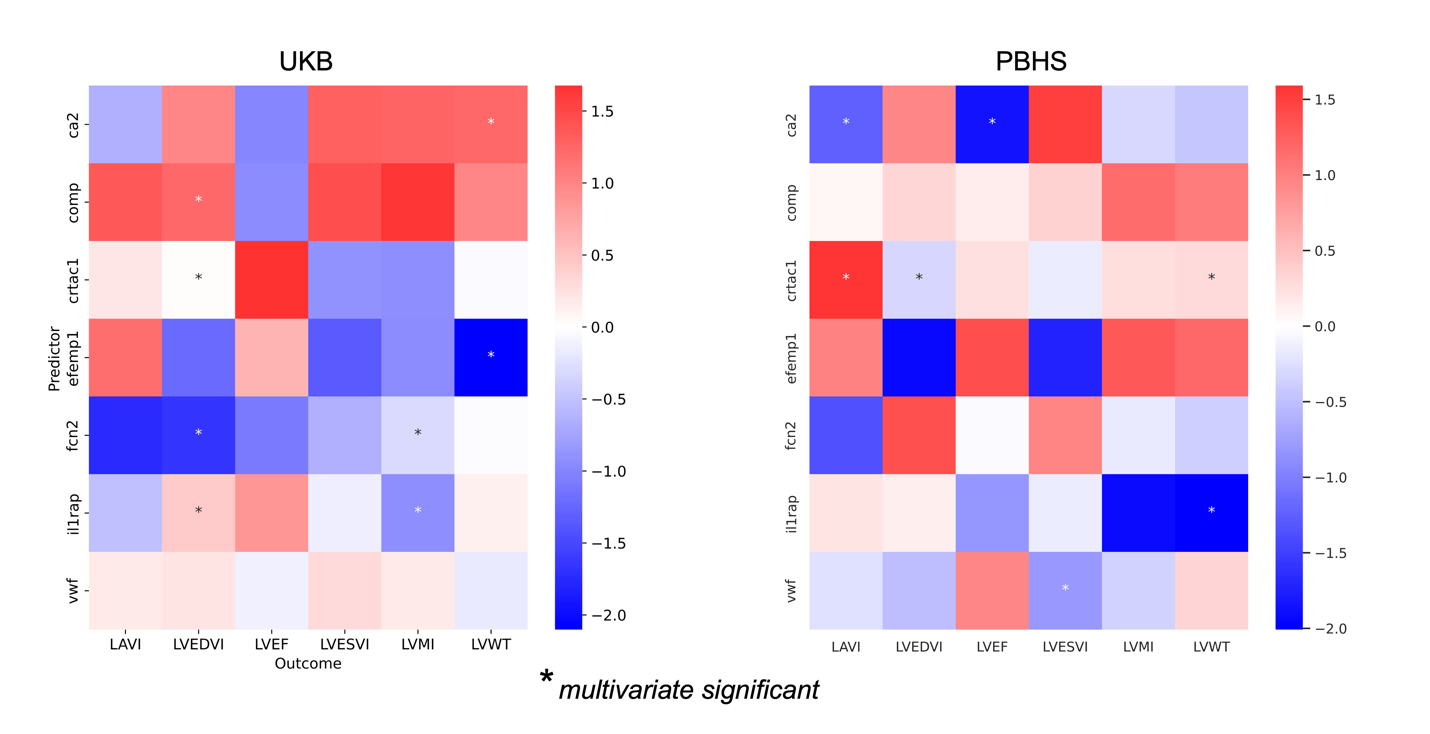

**Supplementary Figure 1**. Overlapping associations between proteins and imaging traits in PBHS and UKB. PBHS: Project Baseline Health Study, UKB: UK Biobank.

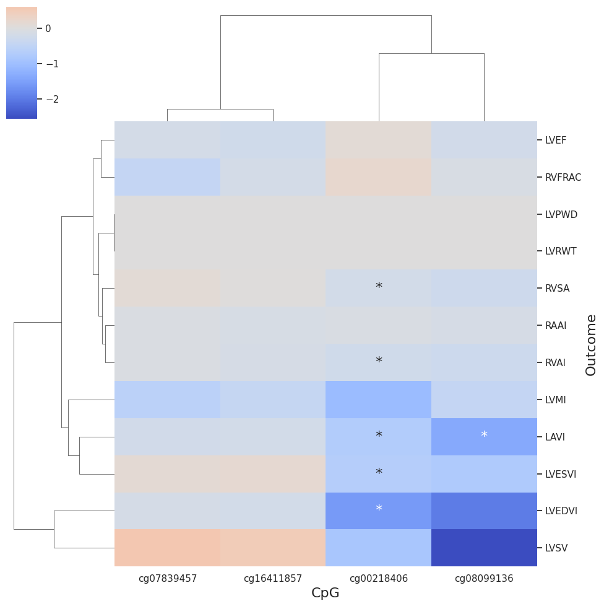

**Supplementary Figure 2.** B2M pQTM (m-value) associations with echocardiographic traits in PBHS. Asterisks (*) represent FDR significant associations, after adjustment for age, sex, smoking status, cell counts. B2M: beta-2 microglobulin, pQTM: protein quantitative trait methylation loci, PBHS: Project Baseline Health Study, FDR: false discovery rate.
